# Estimating the impact of mandatory menu calorie labelling policy and sugar-sweetened beverage taxes on adult obesity prevalence and cardiovascular mortality in two European countries: a simulation modelling study

**DOI:** 10.1101/2024.11.18.24317496

**Authors:** I Gusti Ngurah Edi Putra, Martin O’Flaherty, Karl M. F. Emmert-Fees, Maria Salve Vasquez, Rebecca Evans, Annette Peters, Chris Kypridemos, Nicolas Berger, Eric Robinson, Zoé Colombet

**Affiliations:** Department of Public Health, Policy, and Systems, University of Liverpool, Liverpool, United Kingdom; Professorship of Public Health and Prevention, TUM School of Medicine and Health, Technical University of Munich, Munich, Germany; Department of Epidemiology and Public Health, Sciensano (Scientific Institute of Public Health), Brussels, Belgium; Department of Psychology, University of Liverpool, Liverpool, United Kingdom; Institute of Epidemiology, Helmholtz Zentrum München, Research Center for Environmental Health (GmbH), Neuherberg, Germany; Chair of Epidemiology, Institute for Medical Information Processing, Biometry and Epidemiology (IBE), Faculty of Medicine, LMU Munich, Munich, Germany; German Centre for Diabetes Research (DZD), partner site: Munich-Neuherberg, Germany

**Keywords:** simulation modelling, policy evaluation, food policies, public health policies, Europe

## Abstract

**Background:** Implementing population-based policies such as mandatory menu calorie labelling in out-of-home food businesses and sugar-sweetened beverage (SSB) taxes are promising approaches to improve population health. We aimed to estimate and compare the likely impacts of menu calorie labelling and SSB taxes on reducing obesity prevalence, cardiovascular disease (CVD) mortality, and socioeconomic-related equitable impacts, in two European countries (Belgium and Germany).

**Methods:** We used microsimulation models over a 20-year simulation horizon (2022–2041). For both policies, we modelled the impacts through assumed changes in energy intake due to consumer responses and food industry reformulation. Scenarios of partial (in “large” out-of-home businesses; ≥ 250 employees) and full (in all out-of-home businesses) implementation for menu calorie labelling and different tax rates for SSBs (10%, 20%, 30%) were simulated.

**Findings:** Compared to the counterfactual scenario (e.g., without additional policies), assuming policies effects on both consumer and industry behaviour, menu calorie labelling applied to all out-of-home businesses was estimated to reduce obesity prevalence by 3·61 (95% uncertainty interval-UI: [2·78, 4·30]) and 4·28 (95% UI: [3·64, 5·06]) percentage points and prevent 1600 (95% UI: [400, 3800]) and 30000 (95% UI: [10000, 58000]) CVD deaths in Belgium and Germany over 20 years, respectively. The 30% SSB tax was estimated to reduce obesity prevalence by 0·27 (95% UI: [0·17, 0·43]) and 0·27 (95% UI: [0·17, 0·39]) percentage points and postpone 2500 (95% UI: [800, 5200]) and 16000 (95% UI: [7500, 28000]) CVD deaths in Belgium and Germany, respectively. SSB taxation may have socioeconomic-related equitable impacts, while menu calorie labelling may not.

**Interpretation:** The menu calorie labelling and SSB taxation have substantial impacts in reducing obesity prevalence and preventing CVD deaths in Belgium and Germany. Implementing both policies will be important to reduce obesity and related CVD burden.

**Funding:** European Research Council, National Institute of Health and Care Research

**Research in context:** *Evidence before this study:* We searched simulation modelling studies on the impacts of food-related policies in MEDLINE from 1st January 2000 to 30th September 2024 using the search terms ("food polic*" OR "health polic*" OR “fiscal polic*” OR "SSB" OR "sugar" OR "menu label*" OR "calorie label*" OR "energy label*") AND (simulation OR microsimulation) based on titles and abstracts, restricted to human subjects. We identified 602 articles. To date, a few simulation modelling studies estimated the population-level impacts of mandatory menu calorie labelling. In England, implementation of mandatory menu calorie labelling in all out-of-home food businesses were estimated to reduce obesity prevalence by 2·65 percentage points and prevent 9200 cardiovascular (CVD) deaths over 20 years. Two different studies in the US estimated 27646 CVD deaths and 16700 cancer deaths prevented over lifetime. Given these estimated impacts and England having pioneered the implementation of mandatory menu calorie labelling in Europe, this policy is currently be considered for implementation in many other European countries. However, no studies have examined the potential impacts of implementing this policy in other countries in Europe, nor the extent to which it may offer greater benefits compared to other widely implemented policies, such as sugar-sweetened beverage (SSB) taxes. Many European countries have implemented SSB taxes, including Belgium and the UK. While SSB taxes have been found to be effective in reducing CVD burden based on a scoping review summarising a handful of studies using simulation modelling approaches in different countries, there are gaps in the literature on the impacts of this policy compared to other policies. No studies have estimated and compared the impacts of mandatory menu calorie labelling and SSB taxes on reducing obesity prevalence and CVD mortality in European countries.

*Added value of this study:* This study is the first to estimate and compare the impacts of mandatory menu calorie labelling and SSB taxes in two European countries, Belgium and Germany. Our estimates indicate consistent evidence across both countries for greater benefits in reducing obesity prevalence and CVD mortality from implementing menu calorie labelling in all out-of-home food businesses compared to its implementation limited to large out-of-home food businesses only. While higher tax rates on SSBs were estimated to have bigger benefits, the impact on reducing obesity prevalence was estimated to be smaller compared to mandatory menu calorie labelling in both countries. However, the impact of SSB taxes on CVD mortality was projected to be greater than the mandatory calorie labelling in Belgium, but smaller in Germany. More importantly, based on the current evidence used to inform our models, these policies are complementary as they are estimated to impact CVD mortality through different pathways: mandatory menu calorie labelling primarily affects body mass index (BMI), while SSB taxes mainly operate through a direct BMI-independent effect. In terms of equitable impacts, menu calorie labelling may prevent more deaths in high than low education groups, whereas SSB taxation may postpone more deaths in low than high education groups. Implemented together, these policies will result in greater benefits in addressing diet-related diseases.

*Implications of all the available evidence:* Mandatory menu calorie labelling and SSB taxation were estimated to have substantial impacts on reducing obesity prevalence and preventing CVD mortality. The findings inform the policymakers of both countries and emphasise both the need to implement mandatory menu calorie labelling across out-of-home food businesses and apply higher SSBs tax rates to maximise public health impacts.

## Introduction

In Europe, more than half of adults live with overweight or obesity, with over 20% attributed to obesity alone.^1^ Obesity and its associated physical health burden (e.g., non-communicable diseases (NCDs)) is estimated to have substantial economic impacts in many European countries.^2^ Food environments have been shown to play an important role in influencing diets and the subsequent risk of developing obesity.^3,4^ In line with this, the out-of-home food sector is thought to be a key contributor to the obesity epidemic because eating out is now more commonplace, and out-of-home foods and non-alcoholic beverages (hereafter: food) are characterised by being high in energy.^5,6^ Therefore, public health policies targeting the out-of-home food sector are important in addressing obesity and its adverse health impacts without widening current health inequalities.

Calorie labelling, designed to empower consumers to make healthier choices by providing calorie information at the point of purchase when dining out, has been mandatorily implemented for the first time in Europe, in large out-of-home food businesses (i.e., ≥ 250 employees) in England since 2022.^7,8^ This policy has also been implemented in major chain restaurants with 20 or more outlets in the US,^9,10^ and large chain food businesses in some Australian states.^11^ Based on some previous simulation modelling studies,^8–10^ mandatory menu calorie labelling potentially has population-level impacts in reducing obesity prevalence and NCDs through changing consumer behaviour and inducing industry reformulation. In the US, simulation studies indicate that the implementation of menu calorie labelling in major chain restaurants could prevent 27646 cardiovascular (CVD) deaths^9^ and 16700 cancer deaths over the lifetime of the population.^10^ Full implementation of calorie labelling policy in all out-of-home food businesses in England was estimated to potentially reduce obesity prevalence by 2·65 percentage points and prevent 9200 CVD deaths over 20 years without widening health inequalities, whereas at present, the policy is only implemented in large businesses.^8^ Given these projected impacts, mandatory menu calorie labelling should be considered for implementation in other European countries as part of comprehensive prevention efforts targeting the out-of-home food sector alongside other public health policies like sugar-sweetened beverage (SSB) taxes.

SSB taxation has been adopted in many European countries (e.g., Belgium, France, Ireland, and the UK) to decrease SSB consumption by increasing the prices and encourage businesses to reformulate products to reduce sugar content.^12^ This policy was reported to be promising in preventing obesity and NCDs based on simulation modelling studies from many countries, including Germany, the UK, the US, and Australia.^13,14^ However, a review of studies estimating the impacts of SSB taxes based on simulation modelling approaches concluded that there is limited evidence of (i) the equity impacts of SSB tax across SES groups and (ii) the extent to which SSB tax may offer greater benefits in reducing obesity and CVD mortality compared to other policies,^14^ such as the mandatory menu calorie labelling.

There is a dearth of studies estimating and comparing the impacts of different public health policies in improving population health and assessing impacts on health inequalities. Comparative assessments between different public health policies will be important for policymakers to consider multiple evidence-based policy options and prioritise resources for implementation.^14^ Although structural-based policies (e.g., SSB tax) have been hypothesised to be more effective overall and in reducing health inequality than agency-based policies, which rely on individual motivation (e.g., menu calorie labelling),^13,15^ this may not apply widely. For example, calorie labelling policy does not have differential impacts according to the socioeconomic status based on two meta-analyses^16,17^ and does not seem to widen health inequalities according to a simulation modelling study in England.^8^ In the present study, for the first time, the likely overall population-level impacts and effects across SES groups of implementing calorie labelling and SSB tax were estimated and compared in two European countries with large differences in population size, Belgium and Germany, where obesity prevalence is high (22% for each country^1^) and projected to peak at 30% by 2040.^18^ Menu calorie labelling has not yet been implemented in either Belgium or Germany. While Germany currently does not impose a tax on SSBs, Belgium has an SSB tax in place before 2016. There is limited evidence from both countries on the potential impacts of mandatory menu calorie labelling and how much greater these impacts might be compared to SSB taxation. We aimed to estimate and compare the potential health impacts of implementing menu calorie labelling and varying SSB tax rates in countries with different current policy implementations.

## Methods

We extended a previous simulation model of the calorie labelling policy in England^8^ to estimate and compare the impacts of mandatory menu calorie labelling and SSB taxation on obesity prevalence and CVD (coronary heart disease (CHD) and stroke) mortality in Belgium and Germany. The model,^8^ originally adapted from the IMPACT Food Policy Model,^19^ is a dynamic, stochastic, discrete-time, and open-cohort microsimulation. Under alternative policy scenarios compared to their corresponding counterfactuals (see below), the model simulates the subsequent impact of the policies on relevant exposures (i.e., energy, SSB intake), changes in risk factor (body mass index - BMI), and mortality risk throughout individuals’ life course. We simulated the effects of the two policies over 20 years from 2022 to 2041. We simulated the models from 2022, as this is when the calorie labelling policy was mandatorily implemented for the first time in England.^7,8^ We conducted the simulation separately in Belgium and German populations aged 30 to 89 years using a synthetic population that mimics the real demographic characteristics, BMI, energy and SSB intakes, and disease-related mortality trends using national data sources (see Section “Creating synthetic population” in supplementary materials).

Our scenarios for modelling the impacts of mandatory menu calorie labelling policy in Belgium and Germany followed the current implementation of this policy in England that requires out-of-home “large” food businesses (≥ 250 employees) to display calorie information for non-prepacked food and soft drinks.^7,8^ In Belgium and Germany, large businesses represented 3% and 9% of the number of outlets in the out-of-home food sector and 10% and 21% of this sector turnover in 2019 – 2020, respectively.^20^ We estimated the impacts of mandatory menu calorie labelling policy based on two main scenarios: 1) “partial implementation” scenario, which refers to the implementation in large out-of-home food businesses only (3% and 9% for the number of outlets or 10% and 21% for the sector turnover in Belgium and Germany, respectively) and 2) “full implementation” scenario which refers to the implementation in every out-of-home food business (100% for Belgium and Germany) (see Table 1). For SSB taxes, we modelled different scenarios based on reported changes in SSB consumption following the implementation of the SSB taxes reported by a previous meta-analysis^21^ (see Table 1).

**Table 1.** Scenarios and key assumptions.

| **Mandatory menu calorie labelling** |  |
| --- | --- |
| ***Consumer response*** |  |
| Partial implementation | Based on consumer response (7.3%) and compensation (26.5%) with calorie labelling implemented in large food businesses (3% in Belgium and 9% in Germany) |
| Full implementation | Based on consumer response (7.3%) and compensation (26.5%) with calorie labelling implemented in all food businesses (100% in both Belgium and Germany) |
| ***Reformulation*** |  |
| Partial implementation (Reformulation) | Based on reformulation (5%) with calorie labelling implemented in large food businesses (3% in Belgium and 9% in Germany) |
| Full implementation (Reformulation) | Based on reformulation (5%) with calorie labelling implemented in all food businesses (100% in both Belgium and Germany) |
| ***Combined*** |  |
| Partial implementation | Based on consumer response (7.3%), compensation (26.5%), and reformulation (5%) with calorie labelling implemented in large food businesses (3% in Belgium and 9% in Germany) |
| Full implementation | Based on consumer response (7.3%), compensation (26.5%), and reformulation (5%) with calorie labelling implemented in all food businesses (100% in both Belgium and Germany) |
| **SSB taxes** |  |
| ***Consumer response*** |  |
| 10% tax | Equivalent decrease in SSB consumption due to a 10% increase in price based on a pass-through rate (82%) and a demand price elasticity (–1.59) |
| 20% tax | Equivalent decrease in SSB consumption due to a 20% increase in price based on a pass-through rate (82%) and a demand price elasticity (–1.59) |
| 30% tax | Equivalent decrease in SSB consumption due to a 30% increase in price based on a pass-through rate (82%) and a demand price elasticity (–1.59) |
| ***Reformulation*** |  |
| 30% decrease in sugar | 30% lower sugar content due to industry reformulation, independent of changes in consumer behaviour |
| ***Combined*** |  |
| Combination of consumer response and reformulation | Each of the scenarios of consumer response above combined with reformulation |

For both policies, we compared each scenario with a corresponding counterfactual “baseline” scenario that refers to the current situation or legislation. In Belgium and Germany, “no intervention” served as the counterfactual scenario for modelling mandatory menu calorie labelling as this policy has not yet been implemented. While there is no SSB tax in Germany, Belgium enacted volumetric SSB taxes of €0·03/L before 2016, €0·07/L from 2016, and €0·12/L from 2018.^22^ Our counterfactual scenarios for modelling SSB taxes were “no policy” for Germany and an implemented “SSB tax of €0·03/L” for Belgium because we used SSB consumption data in 2014 (see Section “SSB tax” in supplementary materials).

### Menu calorie labelling effects

We estimated the impact of mandatory menu calorie labelling on energy intake through 1) consumer response (i.e., customers opt for lower-calorie options) and 2) retailer response (i.e., food reformulation of out-of-home retailers) based on two main scenarios: partial and full implementations (e.g., as in ^8^) (see Table 1, Figure 1).

**Figure 1.**
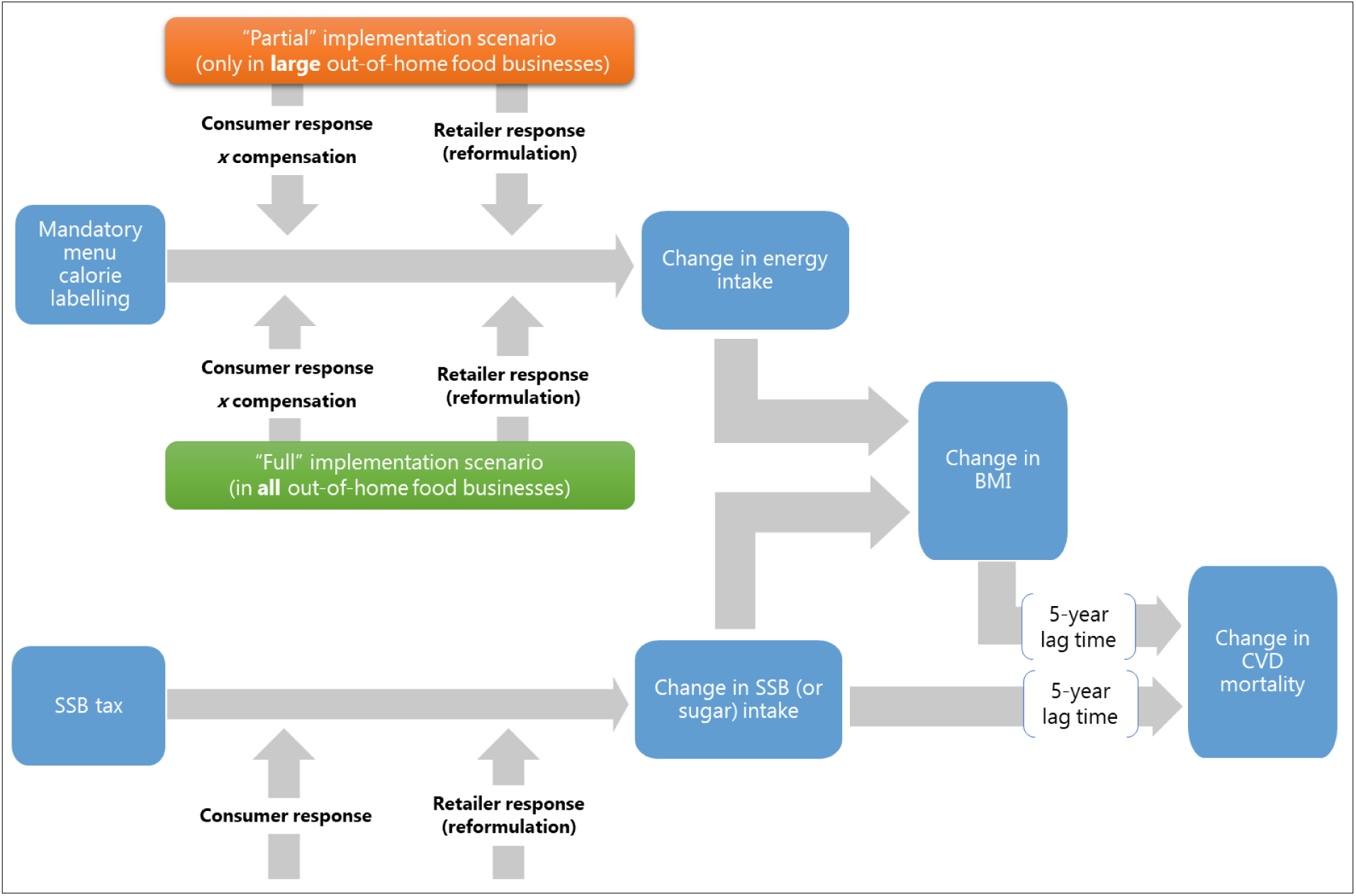
Logic diagram of the impacts of mandatory menu calorie labelling and SSB taxes on CVD mortality (adapted from Colombet et al. ^8^)

#### Effect on consumer response

Following two simulation studies in the US^9,10^ using the findings from a meta-analysis by Shangguan et al.,^23^ we assumed that exposure to calorie labelling would reduce calories intake by 7·3% (95% CI: [−10·1%, −4·4%]) for each out-of-home meal. This effect is similar to a reduction of 47 kcal (95% CI: [−78; −15]) or 7% relative to the average baseline calories purchased (675 kcal) reported in a Cochrane meta-analysis by Crockett et al.^24^ (see Section “Mandatory menu calorie labelling” in supplementary materials). We considered a possible calorie compensation of 26·5% (averaging estimates from two meta-analyses at 42%^25^ and 11%^26^) throughout the day as individuals may consume additional food due to fewer out-of-home calories consumed. Sensitivity analyses with 11% and 42% compensation were conducted. We assumed no differences in the effects of calorie labelling across sociodemographic characteristics following the current literature.^16,17^

#### Reformulation effect

Similar to previous simulation studies,^9,10^ we assumed that calorie labelling would lead to a reduction of 5% in menu options offered by the food businesses. This is based on empirical data of reformulation observed in US chain restaurants.^10^ We assumed the effect was consistent across different menu items due to an absence of contradictory evidence.

### SSB tax effects

We also estimated the impact of the SSB taxes through consumer response and reformulation (see Table 1, Figure 1), even though these two pathways were derived from different SSB tax designs (ad valorem tax and tiered tax, respectively).

#### Effect on consumer response

We developed scenarios for SSB taxes following the findings from a recent meta-analysis by Andreyeva et al.^21^ Based on a pass-through rate of 82% (95% CI: [66%, 98%]) and a demand price elasticity (i.e., % change in sales or consumption due to % change in price) of −1·59 (95% CI: [−2·11, −1·08]),^21^ we modelled SSB ad valorem taxes of 10%, 20%, and 30% (see Section “SSB tax” in supplementary materials). We assumed no substitution to non-SSBs or untaxed beverages due to increased SSB prices, as shown in a high-quality meta-analysis.^21^ Our scenario of a 20% ad valorem tax on SSBs follows a previous modelling study and is based on scientific recommendations.^13,21,27^ We conducted sensitivity analyses using reductions of 6·7% (95% CI: [−10·4, −3·1%]) and 10·0% (95% CI: [−14·7; −5·0]) due to a 10% increase in SSB price reported by Afhsin et al.^28^ and Teng et al.,^29^ respectively. We assumed a similar effect of SSB taxes across socioeconomic groups due to mixed evidence across study settings.^21^

#### Reformulation effect

We assumed that the SSB tax would lead to a reduction of the sugar content of liable beverages by 30% (e.g., as in ^13^) based on the observed effect of the soft drinks industry levy (SDIL) in the UK reported by two studies: 29% decrease in the sugar content of all SSB products sold,^30^ and a 30% decrease in the sugar volume from soft drinks sold.^31^

### Data sources

We created a synthetic population for both countries (see Section “Creating synthetic population”, Appendix Tables 1, 2, and 3 in supplementary materials). We used population projections from Statbel, the Belgian Statistical Office and projected mortality trends based on the CVD deaths observed from 2012 to 2020. The population projections for Germany were from the German Federal Statistical Office and we projected mortality trends based on the CVD deaths observed from 1991 to 2019. For the exposures (BMI, energy, SSB intakes), we used nationally representative surveys: the National Food Consumption Survey (FCS) for Belgium^32^ and *Kooperative Gesundheitsforschung in der Region Augsburg* (KORA) and *Nationale Verzehrstudie* (NVS) II for Germany.^33,34^ We fitted generalised additive models for location, shape and scale (GAMLSS) models to estimate BMI distribution conditional on year (for Germany only), age, sex and education. Energy and SSB intake distributions were conditional on year, age, sex, education, and BMI (e.g. as in ^13^). We calculated out-of-home energy by multiplying energy intakes with the proportion of out-of-home consumption. Similar approach was used to calculate non-diet SSB intake (see Section “Creating synthetic population” in supplementary materials). All data management and statistical analyses were conducted using R Studio. For code see https://github.com/zoecolombet/MenuEnergyLabelling_code_Europe

**Table 2.**
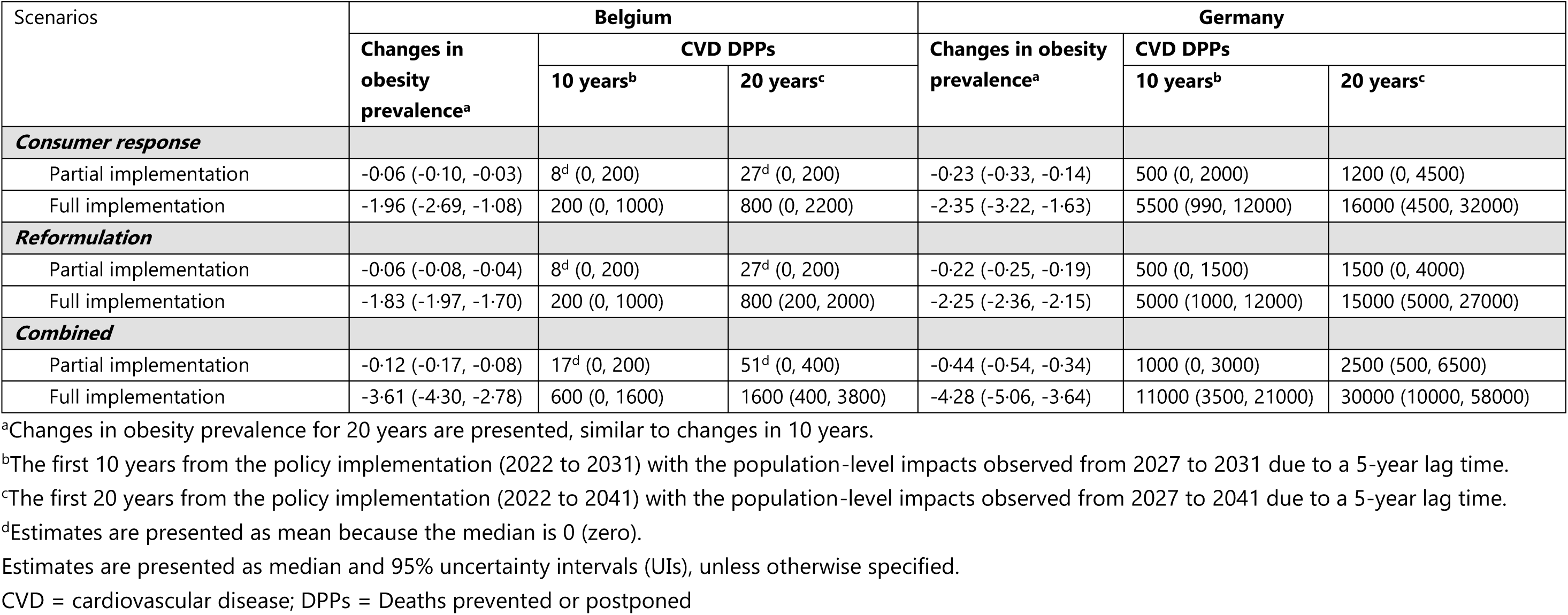
Estimated impacts of mandatory menu calorie labelling on changes in obesity prevalence and CVD mortality in Belgium and Germany (2022 – 2041).

| Scenarios | Belgium |  |  | Germany |  |  |
| --- | --- | --- | --- | --- | --- | --- |
|  | Changes in obesity prevalence<sup>a</sup> | CVD DPPs |  | Changes in obesity prevalence<sup>a</sup> | CVD DPPs |  |
|  |  | 10 years<sup>b</sup> | 20 years<sup>c</sup> |  | 10 years<sup>b</sup> | 20 years<sup>c</sup> |
| ***Consumer response*** |  |  |  |  |  |  |
| Partial implementation | -0.06 (-0.10, -0.03) | 8<sup>d</sup> (0, 200) | 27<sup>d</sup> (0, 200) | -0.23 (-0.33, -0.14) | 500 (0, 2000) | 1200 (0, 4500) |
| Full implementation | -1.96 (-2.69, -1.08) | 200 (0, 1000) | 800 (0, 2200) | -2.35 (-3.22, -1.63) | 5500 (990, 12000) | 16000 (4500, 32000) |
| ***Reformulation*** |  |  |  |  |  |  |
| Partial implementation | -0.06 (-0.08, -0.04) | 8<sup>d</sup> (0, 200) | 27<sup>d</sup> (0, 200) | -0.22 (-0.25, -0.19) | 500 (0, 1500) | 1500 (0, 4000) |
| Full implementation | -1.83 (-1.97, -1.70) | 200 (0, 1000) | 800 (200, 2000) | -2.25 (-2.36, -2.15) | 5000 (1000, 12000) | 15000 (5000, 27000) |
| ***Combined*** |  |  |  |  |  |  |
| Partial implementation | -0.12 (-0.17, -0.08) | 17<sup>d</sup> (0, 200) | 51<sup>d</sup> (0, 400) | -0.44 (-0.54, -0.34) | 1000 (0, 3000) | 2500 (500, 6500) |
| Full implementation | -3.61 (-4.30, -2.78) | 600 (0, 1600) | 1600 (400, 3800) | -4.28 (-5.06, -3.64) | 11000 (3500, 21000) | 30000 (10000, 58000) |

<sup>a</sup>Changes in obesity prevalence for 20 years are presented, similar to changes in 10 years.
<sup>b</sup>The first 10 years from the policy implementation (2022 to 2031) with the population-level impacts observed from 2027 to 2031 due to a 5-year lag time.
<sup>c</sup>The first 20 years from the policy implementation (2022 to 2041) with the population-level impacts observed from 2027 to 2041 due to a 5-year lag time.
<sup>d</sup>Estimates are presented as mean because the median is 0 (zero).
Estimates are presented as median and 95% uncertainty intervals (UIs), unless otherwise specified.
CVD = cardiovascular disease; DPPs = Deaths prevented or postponed

### Model engine

We modified a framework for simulation modelling of mandatory menu calorie labelling in England.^8^ We hypothesised that mandatory menu calorie labelling (partial or full) implementation would immediately reduce the population’s out-of-home energy intake which has further impacts on changes in CVD mortality risk through a reduction in BMI (see Figure 1, and section “Estimating the effect of change in energy and SSB intake on BMI and CVD mortality” in supplementary materials). Changes in energy intake were computed by subtracting post-intervention energy from baseline energy intake every year. The Christiansen & Garby prediction formula^35^ guided how the changes in energy intake were transformed into equivalent changes in body weight before being converted into BMI changes (see Section “Estimating the effect of change in energy intake on BMI” in supplementary materials). Using BMI changes, we estimated subsequent change (or reduction) in obesity prevalence and the 5-year lag-time changes (e.g., as in ^36^) in CVD mortality risk. Based on this information, new mortality rates and the number of deaths were projected. We also adopted calorie labelling framework for estimating the impacts of SSB taxes. The effect of SSB taxes on BMI informed changes in obesity prevalence. For the impacts of SSB taxes on CVD mortality, we estimated the simultaneous effects of SSB intake through two pathways: (i) changes in BMI (or BMI-mediated pathway; indirect effect) and (ii) without BMI pathway (direct effect) (see Figure 1 and Section “Estimating the effect of change in SSB intake on BMI and CVD mortality” in supplementary materials). We assumed that SSB intake has immediate effect on BMI, and we followed menu calorie labelling framework in modelling the subsequent effect on BMI changes on CVD mortality risk. For the direct pathway, we assumed the same 5-year lag time for the effect of SSB intake on CVD mortality risk. We also reported the indirect effect of SSB taxes on CVD mortality through BMI as part of the sensitivity analyses. Because we assumed a 5-year lag time, the policies impact the population from 2027 up to the simulation period to 2041.

### Model outputs

For every scenario in each policy, changes in obesity prevalence and the total number of CVD deaths prevented or postponed (DPPs) were simulated in adults aged 30-89 years separately in Belgium and Germany. To assess the equitable impacts of both these policies based on socioeconomic status, we presented the outputs stratified by educational level and compared the DPP rates between low and high education groups. We presented the findings to two significant digits for DPPs and to two decimal places for changes in prevalence.

### Estimating model uncertainty

We ran 200 iterations using a Monte Carlo method to obtain the uncertainty of model parameters and then reported the results as median values and 95% uncertainty intervals (UIs). See “Estimating model uncertainty” in supplementary materials for detailed information on possible sources of uncertainty.

## Results

### Mandatory menu calorie labelling

Table 2 presents the estimated impacts of mandatory menu calorie labelling on obesity prevalence and preventing or postponing CVD deaths in Belgium and Germany. In both countries, estimated impacts through consumer response were similar to retailer reformulation. We estimated higher impacts on obesity prevalence and CVD mortality for Germany than Belgium when the full scenario of mandatory menu calorie labelling was implemented.

In Belgium, the partial implementation (3% of the total number of outlets) and considering consumer response and reformulation, was estimated to reduce obesity prevalence by 0·12 percentage points (absolute, 95% UI: [0·08, 0·17]) and prevent 51 CVD deaths (95% UI: [0, 400]) over 20 years. However, the full implementation was estimated to have markedly larger impacts on reductions in obesity prevalence (3·61 percentage points; 95% UI: [2·78, 4·30]) and CVD mortality (1600 DPPs; 95% UI: [400, 3800]); which is around 0·90% (95% UI: [0·24, 1·88]) of the total expected CVD deaths.

In Germany, the partial implementation (9% of the total number of outlets) accounting for both consumer response and reformulation would result in a 0·44 percentage-point decline (95% UI: [0·34, 0·54]) in obesity prevalence and 2500 CVD DPPs (95% UI: [500, 6500]) over 20 years. The full implementation in Germany was estimated to reduce obesity prevalence by 4·28 (95% UI: [3·64, 5·06]) percentage points. The reduction in CVD mortality was estimated to be 12 times more than that achieved by implementing this policy in large out-of-home sector businesses only: 30000 CVD deaths (95% UI: [10000, 58000]), around 1·14% (95% UI: [0·51, 1·87]) of the total expected CVD deaths.

Under the full mandatory menu calorie labelling scenario in both countries, we estimated greater changes in obesity prevalence among low and middle education groups than in high education one (Table 3). Comparing rates of CVD DPPs per 100,000 population between low and high education groups, we estimated ratios of 0.86 and 0.78 for Belgium and Germany, respectively (Appendix Table 4). This indicates that menu calorie labelling implemented in all out-of-home businesses may prevent fewer CVD deaths in low than high education groups in both countries.

**Table 3.** Estimated impacts of mandatory menu calorie labelling accounting for combined consumer response and reformulation on obesity prevalence and CVD mortality by educational level in Belgium and Germany (2022 – 2041)

| Scenarios | Belgium |  | Germany |  |
| --- | --- | --- | --- | --- |
|  | Obesity prevalence | CVD deaths | Obesity prevalence | CVD deaths |
| ***Counterfactual*** | ***Predicted*** | ***Predicted*** | ***Predicted*** | ***Predicted*** |
| Low education | 33.54 (32.72, 34.29) | 82000 (48000, 130000) | 26.01 (25.38, 26.73) | 300000 (200000, 420000) |
| Middle education | 32.24 (31.44, 32.97) | 52000 (31000, 76000) | 20.61 (20.27, 20.87) | 1500000 (960000, 2200000) |
| High education | 18.79 (18.11, 19.42) | 37000 (22000, 61000) | 14.80 (14.48, 15.09) | 790000 (520000, 1100000) |
| ***Partial implementation*** | ***Changes*** | ***DPPs*** | ***Changes*** | ***DPPs*** |
| Low education | -0.12 (-0.20, -0.07) | 25<sup>a</sup> (0, 200) | -0.51 (-0.67, -0.39) | 0 (0, 1000) |
| Middle education | -0.13 (-0.21, -0.09) | 17<sup>a</sup> (0, 200) | -0.43 (-0.54, -0.34) | 1500 (0, 4500) |
| High education | -0.10 (-0.15, -0.05) | 9<sup>a</sup> (0, 200) | -0.38 (-0.48, -0.28) | 500 (0, 3000) |
| ***Full implementation*** | ***Changes*** | ***DPPs*** | ***Changes*** | ***DPPs*** |
| Low education | -3.70 (-4.54, -2.85) | 600 (0, 1600) | -5.05 (-6.13, -4.20) | 2800 (500, 6500) |
| Middle education | -4.11 (-4.85, -3.13) | 600 (0, 1400) | -4.23 (-5.03, -3.60) | 16000 (5000, 32000) |
| High education | -2.96 (-3.53, -2.37) | 400 (0, 1200) | -3.58 (-4.23, -3.03) | 10000 (2500, 21000) |

Estimates are presented for 20 years from the policy implementation (2022 to 2041) with the population-level impacts observed from 2027 to 2041 due to a 5- year lag time.
<sup>a</sup>Estimates are presented as mean because the median is 0 (zero).
Estimates are presented as median and 95% uncertainty intervals (UIs), unless otherwise specified.
CVD = cardiovascular disease; DPPs = Deaths prevented or postponed

**Table 4.** Estimated impacts of SSB tax on changes in obesity prevalence and CVD mortality in Belgium and Germany (2022 – 2041).

| Scenarios | Belgium |  |  | Germany |  |  |
| --- | --- | --- | --- | --- | --- | --- |
|  | Changes in obesity prevalence<sup>a</sup> | CVD DPPs |  | Changes in obesity prevalence<sup>a</sup> | CVD DPPs |  |
|  |  | 10 years<sup>b</sup> | 20 years<sup>c</sup> |  | 10 years<sup>b</sup> | 20 years<sup>c</sup> |
| **Consumer response** |  |  |  |  |  |  |
| 10% tax | -0.06 (-0.11, -0.03) | 200 (0, 800) | 600 (0, 2000) | -0.06 (-0.10, -0.03) | 1800 (0, 4500) | 4500 (1000, 9500) |
| 20% tax | -0.12 (-0.23, -0.07) | 400 (0, 1400) | 1200 (200, 2800) | -0.12 (-0.20, -0.06) | 3500 (990, 6500) | 8500 (3000, 16000) |
| 30% tax | -0.18 (-0.34, -0.10) | 600 (0, 1600) | 1700 (400, 4200) | -0.18 (-0.30, -0.10) | 5000 (1500, 9500) | 12000 (4500, 22000) |
| **Reformulation** |  |  |  |  |  |  |
| 30% decrease in sugar | -0.14 (-0.22, -0.09) | 400 (0, 1400) | 1200 (400, 3600) | -0.14 (-0.20, -0.09) | 4000 (1000, 7000) | 10000 (3500, 18000) |
| **Combined** |  |  |  |  |  |  |
| 10% tax | -0.19 (-0.30, -0.12) | 600 (0, 1600) | 1800 (600, 4200) | -0.18 (-0.27, -0.12) | 5000 (1500, 9000) | 12000 (4500, 22000) |
| 20% tax | -0.23 (-0.37, -0.15) | 600 (0, 2000) | 2000 (800, 4800) | -0.23 (-0.32, -0.14) | 5500 (1500, 11000) | 15000 (6500, 26000) |
| 30% tax | -0.27 (-0.43, -0.17) | 800 (0, 2200) | 2500 (800, 5200) | -0.27 (-0.39, -0.17) | 6500 (2000, 12000) | 16000 (7500, 28000) |

<sup>a</sup>Changes in obesity prevalence for 20 years are presented, similar to changes in 10 years.
<sup>b</sup>The first 10 years from the policy implementation (2022 to 2031) with the population-level impacts observed from 2027 to 2031 due to a 5-year lag time.
<sup>c</sup>The first 20 years from the policy implementation (2022 to 2041) with the population-level impacts observed from 2027 to 2041 due to a 5-year lag time.
<sup>d</sup>Estimates are presented as mean because the median is 0 (zero).
Estimates are presented as median and 95% uncertainty intervals (UIs), unless otherwise specified.
CVD = cardiovascular disease; DPPs = Deaths prevented or postponed

Our sensitivity analyses produced comparable findings. Larger impacts were estimated when using minimum (11%) than maximum (42%) compensation (Appendix Table 5) and using turnover than the proportion of outlets for the partial implementation scenario (Appendix Table 6).

**Table 5.** Estimated impacts of the SSB tax accounting for combined consumer response and reformulation on obesity prevalence and CVD mortality by educational level in Belgium and Germany (2022 – 2041)

| Scenarios | Belgium |  | Germany |  |
| --- | --- | --- | --- | --- |
|  | Obesity prevalence | CVD deaths | Obesity prevalence | CVD deaths |
| **Counterfactual** | **Predicted** | **Predicted** | **Predicted** | **Predicted** |
| Low education | 33·54 (32·72, 34·29) | 82000 (48000, 130000) | 26·01 (25·38, 26·73) | 300000 (200000, 420000) |
| Middle education | 32·24 (31·44, 32·97) | 52000 (31000, 76000) | 20·61 (20·27, 20·87) | 1500000 (960000, 2200000) |
| High education | 18·79 (18·11, 19·42) | 37000 (22000, 61000) | 14·80 (14·48, 15·09) | 790000 (520000, 1100000) |
| **10% tax** | **Changes** | **DPPs** | **Changes** | **DPPs** |
| Low education | -0·23 (-0·37, -0·13) | 800 (200, 2200) | -0·27 (-0·42, -0·16) | 2200 (500, 5500) |
| Middle education | -0·24 (-0·39, -0·15) | 800 (0, 1800) | -0·16 (-0·23, -0·10) | 7000 (2000, 13000) |
| High education | -0·09 (-0·14, -0·03) | 200 (0, 800) | -0·12 (-0·18, -0·07) | 3500 (990, 7000) |
| **20% tax** | **Changes** | **DPPs** | **Changes** | **DPPs** |
| Low education | -0·28 (-0·46, -0·17) | 1000 (200, 2600) | -0·34 (-0·50, -0·19) | 2500 (990, 6000) |
| Middle education | -0·30 (-0·47, -0·18) | 800 (200, 2000) | -0·19 (-0·28, -0·12) | 8000 (3000, 15000) |
| High education | -0·11 (-0·18, -0·05) | 200 (0, 1000) | -0·15 (-0·22, -0·09) | 3500 (1000, 8000) |
| **30% tax** | **Changes** | **DPPs** | **Changes** | **DPPs** |
| Low education | -0·34 (-0·54, -0·20) | 1200 (200, 2800) | -0·39 (-0·62, -0·24) | 3000 (1000, 6500) |
| Middle education | -0·35 (-0·57, -0·21) | 1000 (200, 2200) | -0·23 (-0·34, -0·15) | 8800 (3000, 16000) |
| High education | -0·13 (-0·21, -0·06) | 200 (0, 1000) | -0·17 (-0·26, -0·10) | 4000 (1500, 8500) |

Estimates are presented for 20 years from the policy implementation (2022 to 2041) with the population-level impacts observed from 2027 to 2041 due to a 5- year lag time.
Estimates are presented as median and 95% uncertainty intervals (UIs), unless otherwise specified.
CVD = cardiovascular disease; DPPs = Deaths prevented or postponed

### SSB tax

The likely population-level impacts of SSB tax on obesity prevalence and CVD mortality through consumer response increased in line with the higher tax rates implemented (Table 4). The reformulation effect (sugar content reduced by 30%) alone was estimated to have similar impacts than implementing a 20% SSB tax. The estimated changes in obesity prevalence resulting from SSB taxes were similar in Belgium and Germany.

In Belgium, consumer response to a 20% and 30% tax rates would reduce obesity prevalence by 0·412 (95% UI: [0·07, 0·23]) and 0·18 (95% UI: [0·10, 0·34]) percentage points and prevent 1200 (95% UI: [200, 2800]) and 1700 (95% UI: [400, 4200]) CVD deaths over two decades. Reformulation alone was estimated to result in a 0·14 percentage point (95% UI: [0·09, 0·22]) decline in obesity prevalence and 1200 CVD DPPs (95% UI: [400, 3600]). Combining both consumer response and reformulation would result in bigger estimated impacts. For example, a 30% tax rate combined with reformulation was estimated to decrease obesity prevalence by 0·27 (95% UI: [0·17, 0·43]) percentage points and postpone 2500 deaths (95% UI: [800, 5200]) or around 1.46% (95% UI: [0.54%, 3.07%]) of the expected CVD deaths.

In Germany, consumer response to the implementation of SSB taxes would yield a 0·12-percentage point (95% UI: [0·06, 0·20]) decline in obesity prevalence and 8500 (95% UI: [3000, 16000]) CVD DPPs for the 20% SSB tax and a 0·18-percentage point (95% UI: [0·10, 0·30]) decline in obesity prevalence and 12000 CVD DPPs (95% UI: [4500, 22000] for the 30% SSB tax. The reformulation alone would decline obesity prevalence by 0·14 percentage points (95% UI: [0.09, 0.20]) and reduce CVD deaths by 10000 (95% UI: [3500, 18000]). We estimated bigger impacts from combining consumer response and reformulation with a decline in obesity prevalence by 0·27 percentage points (95% UI: 0·17, 0·39) and a reduction in CVD deaths by 16000 (95% UI: 7500, 28000) (around 0.62% (95% UI [0.27%, 0.97%]) of the predicted deaths) if a 30% SSB tax would be implemented.

We estimated greater changes in obesity prevalence in low than in higher education groups in Belgium and Germany (Table 5). We compared rates of CVD DPPs of implementing a 30% SSB tax considering both consumer response and reformulation per 100,000 population between low and high education groups. We estimated ratios of 3.31 and 2.00 for Belgium and Germany, respectively, and the probability of the ratios > 1 was higher than 50% (Appendix Table 4). This indicates that the policy may prevent more CVD deaths in low than high education groups in both countries.

Sensitivity analyses using effect sizes from different meta-analyses showed similar findings to the primary analyses of the same 10% SSB tax rate (Appendix Table 7). We found small impacts when the effect of SSB taxes on a reduction in CVD mortality was only estimated through changes in BMI (Appendix Table 8).

## Discussion

To inform future food policy in Europe, we modelled the likely population impacts of implementing mandatory menu calorie labelling and SSB taxes on obesity prevalence and CVD mortality in two European countries (2022 – 2041). In both countries, we estimated the impact of menu calorie labelling on obesity prevalence to be greater than SSB taxes when implemented in every out-of-home business. However, implementing menu calorie labelling in all out-of-home businesses is estimated to postpone or prevent more CVD deaths than the highest SSB tax rate (30%) in Germany (1·14% vs 0·62% of the total expected CVD deaths) but not in Belgium (0·90% vs 1·46%). Under the assumption that the policies have the same effect across ages, sexes, and SES groups,^8^ we estimated that SSB taxation may have equitable impacts as the policy tend to prevent more CVD deaths in low than high education groups. However, menu calorie labelling may not have equitable impacts as more CVD DPPs were estimated in high than low education groups.

Mandatory menu calorie labelling would have a higher impact across the studied countries if the policy were implemented for all out-of-home businesses, rather than just large businesses, as is currently the case in England.^8^ Our findings are consistent with a previous simulation modelling in England,^8^ suggesting large impacts of implementing mandatory menu calorie labelling in all out-of-home sectors with obesity prevalence reduced by 2·65 percentage points. This study also estimated 9200 CVD DPPs or around 1·10% (95% UI 0·71–1·60) relative to the expected CVD deaths,^8^ which is similar to our estimates in Belgium (0·90%) and Germany (1·14%).^8^ Our results are also consistent with previous modelling in the US.^9^ For example, our full scenario without reformulation (a compensation level of 26.5%) would result in 16000 CVD DPPs in Germany compared to 27646 CVD DPPs in the US (a higher compensation of 50% with much larger population size).^9^ In addition, our research echoes their finding that adding the reformulation doubles the mortality benefits.^9^

Our findings for SSB taxation are also similar to those of previous simulation modelling in Germany.^13^ Under the same scenario of 20% SSB tax without reformulation assumed, our estimates of 8500 CVD DPPs are half of 17000 all-cause DPPs (combined CVD and non-CVD deaths) reported by the previous modelling (findings on CVD deaths only are not presented).^13^ A handful of studies have simulated the impact of sugary drink policies across different study contexts and reported consistent results.^14^ For example, a study from the US with a larger population size estimated 31000 CVD DPPs in the next 15 years from implementing a 10% SSB tax.^19^ It is important to note that the likely impacts on CVD mortality estimated through BMI only (Appendix Table 8) were similarly modest in size compared to the previous modelling estimates.^13^ In line with this, the benefits of reducing obesity prevalence were much smaller than the estimates for mandatory calorie labelling, indicating that the SSB tax may largely impact CVD mortality through a pathway not involving changes in BMI as discussed in a previous simulation modelling study.^13^

We estimated that mandatory menu calorie labelling may have greater population-level impacts than SSB tax in reducing obesity prevalence in both countries, and in preventing CVD deaths in Germany but not in Belgium. Greater impacts of menu calorie labelling on reducing obesity prevalence may be because energy intake from out-of-home has more direct and substantial impacts on weight gain, particularly due to larger portion sizes (volume) and high in fat and overall calorie content.^5,6,37,38^ In line with this, a study of UK Biobank participants reported that BMI has stronger associations with total energy and energy from fat than sugar.^39^ The greater CVD mortality-related benefits of SSB taxes compared to menu calorie labelling in Belgium, but not in Germany, may be explained by higher SSB intake in Belgium (see Appendix Table 3). Importantly, the evidence used to inform our model suggests that these policies may impact CVD mortality through different pathways that involve changes in BMI for mandatory menu calorie labelling and other BMI-independent mechanisms for SSB taxes. Therefore, the policies are complementary, and implementing both as part of the public health efforts in addressing diet-related diseases will yield greater benefits in both countries. It is important to note that menu calorie labelling may not have equitable impacts in Belgium and Germany as the policy tends to postpone more CVD deaths in high than low education groups. This can be explained by our estimated exposure data, which shows higher out-of-home energy intake in high than low education groups (see Appendix Table 3). However, the estimated out-of-energy intake in our models was calculated by sex and age groups only due to the data unavailability by education. Future modelling studies in other settings are warranted to provide more insights on the relevance of this assumption.

The present study has some strengths. To our knowledge, this is the first comparison of likely health impact of mandatory menu calorie labelling and SSB taxation in a European setting. Our findings are particularly robust as we used a validated model that previously has been exercised to estimate the calorie labelling in England and other dietary specific policies in the US.^8,19^ Scenarios were informed from current policy practices (e.g., mandatory menu calorie labelling in England). Our estimates are supported by rigorous sensitivity analyses that account for uncertainties in modelling assumptions and are consistent with findings from previous studies,^8,9,13,14,19^ increasing confidence in the results. Two different policies were examined using the same model framework, addressing the current gaps highlighted by a scoping review^14^ on a dearth of evidence on the impacts of SSB policy compared to other policies.

The present study also has some limitations. First, we used a proportional effect estimate (7·3% reduction)^23^ for the main analysis. Even though this estimate has been used in previous simulation modelling studies^9,10^ and similar to findings from another meta-analysis others may not be transferable to the population in Belgium and Germany as the effect may vary depending on the study context. While there is an absence of evidence for Belgium and Germany, observational studies showed that implementation of menu calorie labelling in a single fast food chain across three US states was associated with a purchase reduction per customer of 82 calories,^40^ but not such reduction in calories per transaction was observed in England from a recent pre-post comparison study.^41^ This limitation applies to the reformulation effect of menu calorie labelling and the effect estimates (consumer response, reformulation) used for simulating the effects of SSB tax. Evidence based on empirical impacts of the policies in both countries would improve the precision of modelling the long-term policy impacts. we assumed the effects of policies remain stable throughout the simulation period due to the absence of contrasting evidence. However, this is not always the case as the effect may change (e.g., decrease due to habituation to information, or increase due to increased awareness and policy campaign) over time.^8^ We also did not consider cumulative effects of out-of-home energy and SSB intakes over the life course

Our exposures (i.e., BMI, energy, SSB intakes) were based on the most recent available representative surveys in 2014 or earlier, and we assumed the patterns have continue since then. Similarly, the proportions of out-of-home intakes were derived from studies conducted in early 2000s, assuming no subsequent changes by age groups and sex. While these sources are the best available, dietary habits may change in respond to the COVID-19 pandemic^42^ and recent economic downturns.^43^ We assume that these limitations would be more likely to underestimate rather than overestimate policy impacts. For example, in the context of the menu calorie labelling policy, eating out may now be more common reflected by the increased numbers of out-of-home food retailers in the last 20 years in both countries.^20^ For Belgium, we used the €0.03/L tax (before 2016) as the counterfactual scenario because we used SSB consumption data in 2014. SSB intake may have decreased due to a new tax of 0.12/L implemented in 2018. We assumed this price increase, from €0.03/L to €0.12/L, is approximately equivalent to a 10% SSB tax (see Section “SSB tax” in supplementary materials). Therefore, implementing higher SSB taxes (20% or 30%) would result in greater impacts than the current implemented SSB tax rate (€0.12/L).

We modelled the impacts specific to the population aged 30-89 years, and therefore, the results do not capture potential policy benefits related to peak SSB consumption in younger ages,^44^ nor do the results account for changes in obesity from childhood to young adults. Finally, we modelled the exposures conditional on education level and we estimated the policy impacts across these education groups. Consequently, we excluded individuals with no information (“unknown”, “not applicable”) on education level in Belgium and this is a limitation. In addition, we assumed the same price elasticity across all SES groups due to mixed evidence.^21^ However, low SES group may benefit more from SSB taxes as they tend to be more responsive to price increases.^45^

Our findings provide new evidence that implementing mandatory menu calorie labelling across all out-of-home food establishments and applying higher tax rates (e.g., 30%) on SSBs would yield substantial public health benefits by reducing obesity prevalence and preventing CVD deaths. Each of the policies has also been demonstrated to be cost-effective by previous studies.^10,13^ In Belgium, implementing mandatory menu calorie labelling in all out-of-home sectors together with higher tax rates is recommended to maximise public health efforts to tackle diet-related diseases. As neither of the policies has been adopted in Germany, our results emphasise the need for the government to take ambitious steps towards implementing both mandatory menu calorie labelling policy in all-out-home businesses and SSB tax at higher rates for greater public health benefits. More importantly, these policies need to be seen as complementary approaches, and with additional measures across the food system highlighting the fact that no single policy will be enough to significantly reduce the burden of unhealthy diets in populations.

## Conclusion

This study provides the first evidence of population-level benefits of implementing national level mandatory menu calorie labelling policy and SSB taxes in Belgium and Germany. Implementing both policies is needed in order to tackle obesity and CVD burden in both countries.

## Data Availability

We used data from different sources. For Belgium, population projection and census data are available online on the official website of Statbel, the Belgian Statistical Office. Projected mortality data are available upon request from Statbel. Exposure data (BMI, energy and SSB intakes) are from the National Food Consumption Survey, available upon request to the data custodian (Sciensano). For Germany, population projection, census, and mortality data are available online on the German Federal Statistical Office's website. Exposure data are from KORA (https://www.helmholtz-munich.de/en/epi/cohort/kora) and NVS surveys available with the permission from the data custodian. R script to generate the results presented in this study is publicly available at https://github.com/zoecolombet/MenuEnergyLabelling_code_Europe.

## Acknowledgements

ER and ZC were part-funded by the European Research Council (Grant reference: PIDS, 803194). IGNEP is funded by the National Institute of Health and Care Research (NIHR) Development and Skill Enhancement Award (DSE) (Grant reference: NIHR305076). ER and RE are funded by the NIHR Oxford Health Biomedical Research Centre (BRC) (Grant reference: NIHR203316). The views expressed are those of the authors and not those of the funders.

## Authors’ contributions

IGNEP, MO’F, ER, ZC designed the study. IGNEP, CK, ZC developed the model. MO’F, CK, and ZC supervised IGNEP. MSV and KME-F did the GAMLSS fitting for exposure data in Belgium and Germany respectively. NB and AP acquired the survey data for Belgium and Germany, respectively. IGNEP and ZC did the analysis and drafted the initial manuscript. RE and ZC verified the results from the analysis. All the authors contributed to the data interpretation and revised the manuscript draft. All the authors approved and accept responsibility to submit for publication.

