## Supplementary information for "Estimating the impact of mandatory menu calorie labelling policy and sugar-sweetened beverage taxes on adult obesity prevalence and cardiovascular mortality in two European countries: a simulation modelling study"

### Modelling approach

We extended a comparative risk assessment model previously developed to estimate the impacts of mandatory menu calorie labelling in England.[1] The model,[1] originally adapted from the IMPACT Food Policy Model[2] was modified to incorporate dynamic, stochastic, discrete-time, and open-cohort microsimulation for this present study. We used this updated model to estimate the likely population-level impacts of mandatory menu calorie labelling and SSB tax in Belgium and Germany over a 20-year horizon from 2022 to 2041. We selected 2022 as the initial year for the simulation modelling following the year the mandatory menu calorie labelling policy was officially implemented for the first time in England and also Europe.[1,3] The simulation modelling was conducted using R Studio (see https://github.com/zoecolombet/MenuEnergyLabelling_code_Europe for the R script).

### Mandatory menu calorie labelling

#### *Scenarios and coverages*

We compared two main scenarios: 1) "partial implementation" which refers to mandatory menu calorie labelling applied to large out-of-home food businesses only (≥250 employees) following the current implementation of this policy in England,[1,3] and 2) "full implementation" which extends this policy to every out-of-home food business. Both scenarios were compared to a counterfactual "no intervention" (baseline) scenario as this policy has not yet been implemented in Belgium and Germany.

We used the most updated data from Eurostat (European Statistical Office, that provide official and harmonised data for the European Union members)[4] to determine the proportions of large out-of-home food businesses (≥250 employees) in Belgium and Germany. As the number of outlets for different sizes of enterprises (micro, small, medium, large) was not available, we used the average number of outlets (or sites) by sizes of businesses in the UK (1 outlet for micro businesses, 1.08 outlets for small businesses, 2.43 outlets for medium businesses, and 63.16 outlets for large businesses).[5,6] We combined information from Eurostat[4] and the number of outlets by business size in the UK[5,6] to calculate the proportion of out-of-home large businesses in Belgium and Germany. In Belgium, out-of-home large business outlets accounted for 3% of the total outlets and this type of business contributed to 10% of the turnover in 2019.[4] In Germany, large out-of-home businesses represented 9% of the number of food outlets and 21% of the turnover in this sector in 2020.[4]

| Size of business | Belgium (2019) | | Germany (2020) | |
|---|---|---|---|---|
| | Number of outlets | Turnover | Number of outlets | Turnover |
| Micro (<10 employees) | 48,174 (90%) | 10,747 (57%) | 171,103 (70%) | 18,843 (30%) |
| Small (10-49 employees) | 3,040 (6%) | 4,547 (24%) | 43,156 (18%) | 19,738 (32%) |
| Medium (50-249 employees) | 382 (1%) | 1,528 (8%) | 7,681 (3%) | 10,651 (17%) |
| Large (≥250 employees) | 1,516 (3%) | 1,944 (10%) | 21,095 (9%) | 12,811 (21%) |
| **Total** | **53,112 (100%)** | **18,766 (100%)** | **243,035 (100%)** | **62,044 (100%)** |

The "partial implementation" scenario estimated the impact of mandatory menu calorie labelling in large out-of-home food businesses (3% and 9% for Belgium and Germany, respectively). The "full implementation" scenario estimated the likely impact of this policy if it was applied to every out-of-home food business (100%). We assumed that the proportions of different businesses are equivalent to the proportions of out-of-home calories consumed from those businesses (as the coverage of the policy). For example, large businesses account for 3% in Belgium and 9% in Germany, and therefore, we assumed that 3% and 9% of the out-of-home calories consumed are from large businesses in Belgium and Germany, respectively. We opted for the number of businesses over turnover as it better reflects the exposure to menu calorie labelling. However, we conducted a sensitivity analysis for the partial implementation scenario using turnover (10% in Belgium and 21% in Germany).

#### ***Effect of mandatory menu calorie labelling on energy intake***

We modelled the likely impact of mandatory menu calorie labelling policy on energy intake through two pathways: 1) consumer response (i.e., customers opt for healthier or lower-calorie options) and 2) retailer response (i.e., food reformulation of out-of-home retailers) (e.g., as in [1]). For both scenarios ("partial" and "full" implementations), we estimated the impacts through these two separate and combined pathways. We assumed the consumer and reformulation effects were stable over the simulation horizon (e.g., as in [1]).

Consumer response to mandatory menu calorie labelling

To model the impact of menu calorie labelling on energy ordered or consumed, we followed two previous simulation modelling studies in the US[7,8] using an estimate from a meta-analysis of 19 intervention studies and randomized control trials (RCTs) conducted by Shangguan et al.[9] Based on this meta-analysis, exposure to menu calorie labelling led to a reduction in energy intake by a 7.3% (95% CI: [-10.1%, -4.4%]). The effect is similar to findings from a Cochrane meta-analysis of three RCTs by Crockett et al.[10] that estimated a reduction of 47 kcal (95% CI: [-78; -15]) per meal on average. This reduction is equivalent to 7.8% (95% CI: [-13.1%, -2.5%]) assuming an average meal of 600 kcal or 7% relative to the average baseline calories purchased in the included RCTs (675 kcal).[10] We used a relative proportional change in energy intake (7.3%) as we did not have information about the frequency of eating out-of-home in the countries studied. Thus, we assumed that implementing menu calorie labelling in out-of-home sectors would reduce out-of-home energy intake by 7.3% (95% CI: [-10.1%, -4.4%]). This assumption is evenly applied across sociodemographic characteristics, such as age, sex, and socioeconomic position, as current evidence suggests that there are no differences in the policy's effects based on these characteristics.[11,12]

Consumers' reduction in energy intake in out-of-home settings in response to menu calorie labelling may be compensated for by consuming additional meals or products throughout the day.[13,14] Recent systematic reviews indicated levels of compensation of 42%[13] and 11%[14] later in the day after consuming less food (volume) and selecting lower energy-density meals, respectively and we used the average of both compensation levels (26.5%) in our main simulation modelling. Nevertheless, sensitivity analyses with 11 and 42% compensation levels were also conducted.

We assumed that everyone who purchased meals prepared out-of-home would be impacted by the policy. We calculated baseline out-of-home energy intake (in kcal) by multiplying energy intake and the proportion energy consumed from out of home (see section "Out-of-home energy intake"). The effect of calorie labeling, accounting for compensation behaviors, was applied to projected out-of-home energy intake to estimate annual changes in energy intake based on the policy's coverage (partial or full implementation), resulting in the post-implementation (or -intervention) energy intake.

Reformulation effect due to menu calorie labelling

We also followed the US simulation modelling studies on an average reduction of 5% in the calorie content of menu items due to mandatory menu calorie disclosure (reformulation).[7,8] This is based on a reformulation observed in the US chain restaurants following the implementation of menu calorie labelling policy.[8,15-17] The 5% reformulation aligns with the findings from a meta-analysis by Zlatevska et al.,[18] suggesting an average reduction of 15 kcal (95% CI: [-23; -8]) in the calorie content of menu items or approximately 4% relative to average baseline calories of 400. We thus assumed that implementing the menu calorie labelling policy would lead to a 5% decrease in the energy content of the products offered in out-of-home businesses; corresponding to a 5% decrease in energy intake from out-of-home. We multiplied this reformulation-associated calorie reduction by the policy coverage according to the scenarios in each country.

### Sugar sweetened beverage (SSB) tax

***Scenarios and coverages***

Different scenarios based on different effects of SSB taxes on SBB consumption from a meta-analysis was implemented (see "Consumer response to menu calorie labelling" below) in Belgium and Germany. These scenarios were compared to counterfactual (baseline) scenarios reflecting current situation on SSB taxes in each country. In Germany, "no intervention" served as a counterfactual scenario as the SSB tax has not yet been deployed. However, Belgium has implemented a volumetric SSB tax for all non-alcoholic drinks with added sugar of €0.03/L before 2016, €0.07/L from 2016, and €0.12/L from 2018.[19] As baseline SSB intakes came from a 2014 survey (see section "Non-diet SSB intake"), we use the SSB tax in place in 2014 as counterfactual scenario for Belgium (SSB tax of €0.03/L before 2016).

No data was available to determine the percentage increase in SSB prices due to the SSB tax in Belgium. Based on a recent study on SSB taxes in European countries,[19] a SSB tax of €0.07/L in France (introduced in 2012) was equivalent to 7%-10% increase in price. Taking the average percentage of the increase in SSB prices in France (8.5%) and assuming similar SSB prices in Belgium and France, a tax of €0.03/L in Belgium was equal to a 3.6% (€0.03 x 8.5% / €0.07) increase in price and a tax of €0.12/L was equivalent to a 14.6% (€0.12 x 8.5% / €0.07) increase in price (assuming a stable inflation rate). Therefore, the new tax of €0.12/L (since 2018) equates to an 11% increase (from 3.6% to 14.6%) in SSB prices. Using the SSB tax of "€0.03/L" as the counterfactual scenario in Belgium, we modelled the effects of ad valorem taxes of 10%, 20%, and 30%. Even though SSB taxes in Belgium are in the form of a volumetric tax, we assumed the effect of increased price (10%) is similar to ad valorem tax at the same rate (10% SSB tax, excluding pass-through) due to lack of product-level ingredient, volume, and price data (see [20,21]). Thus, based on the counterfactual scenario of  0.03/L, our scenarios of increasing the SSB taxes by 10%, 20%, and 30% in Belgium would be equivalent to increased prices to 13.6% (an increase of 10% + 3.6% from counterfactual; the equivalent of a €0.11/L tax (calculated as 13.6% x €0.03 / 3.6%; **which is close to the current tax at €0.12/L**), 23.6% (or €0.20/L (23.6% x €0.03 / 3.6%)), and 33.6% (€0.28/L (33.6% x €0.03 / 3.6%)), respectively. In both countries, we assumed a 100% coverage of the SSB tax as it applies to all SSBs across all types of businesses and retail stores.

***Effect of SSB tax on SSB intake***

As for menu calorie labelling policy, we modelled the likely impact of the SSB tax policy on intake through two pathways: 1) consumer response (i.e., discouraging SSB consumption) and 2) reformulation (i.e., reducing the sugar content of SSBs). Even though the assumed effects above were derived from different SSB tax designs (ad valorem tax for consumer response, tiered tax for reformulation), we aimed to estimate the likely impacts through different pathways and hypothetical combined pathways. We assumed that the effects of the SSB tax on both consumer response and reformulation remained consistent throughout the simulation period.

<u>Consumer response to SSB tax</u>

We developed our modelling scenarios for the impact of SSB taxes based on an effect reported in a previous meta-analysis by Andreyeva et al.[22] We calculated changes in SSB intakes based on a demand price elasticity (i.e., % change in sales or consumption due to % change in price) of −1.59 (95% CI: [−2.11, −1.08]) and a pass-through rate (i.e., the extent of increase in price passed on to customers) of 82% (95% CI: [66%, 98%] reported in a meta-analysis of 33-41 studies. Using this information, we modelled SSB taxes of 10%, 20%, and 30%.

We also conducted sensitivity analyses using two meta-analyses by Afhsin et al.[23] and Teng et al.[24] to model the effect of a 10% SSB tax. Afhsin et al.[23] reported that a 10% increase in SSB price was associated with a 6.7% reduction in SSB intake (95% CI: [-10.4, -3.1%]) calculated from a meta-analysis of three non-randomised interventions and two prospective cohort studies. In a meta-analysis of 17 pre-post intervention comparisons (the majority used interrupted time series analysis) by Teng et al.,[24] a 10.0% increase in SSB price was associated with a decline in intake and purchases by 10% (95% CI : [-14.7; -5.0]).

We assumed no substitution to non-SSBs or untaxed beverages (e.g., juice, milk) due to an increase in SSB prices as shown in a meta-analysis by Andreyeva et al.[22] The effect was modelled consistently across sociodemographic characteristics due to limited data on the heterogeneous effects of SSB tax in different sub-populations.[22] Everyone consuming non-diet SSBs was assumed to be impacted by the policy. We calculated baseline non-diet SSB consumption by multiplying overall SSB intake (in mL or grams; 1 mL = 1 grams) with the proportion of non-diet SSB intake in Germany. Because almost all the participants from the survey on which the SSB intake was based were consumers of sugary drinks (99-100%) in Belgium, we assumed that all SSB intake was from non-diet SSBs (see section "Non-diet SSB intake"). The effects of different scenarios of SSB taxes were applied to the projected non-diet SSB intake to calculate annual changes in intake and post-intervention SSB intake.

Reformulation effect due to SSB tax

The soft drinks industry levy (SDIL) in the UK has been observed to reduce the sugar content of all SSB products sold by 28.5%[25] or the volume of sugars sold from all soft drinks by 30%.[26] Therefore, we assumed a 30% lower sugar content due to (a tiered) SSB tax independently of change in consumption (e.g., as in [20]). We assumed that one SSB serving of 227.3045 mL (8 oz) contains 20 grams of sugar (e.g. as in [20,27]). Thus, the reformulation would reduce sugar content by 30% (or 6 out of 20 grams), and therefore, a post-intervention SSB serving will have 14 grams of sugar per 227.3045 mL.

### Creating synthetic population

We created a synthetic population of Belgium and a synthetic population of Germany to simulate and estimate the population-level impact of mandatory menu calorie labelling and SSB tax scenarios. Data that we used in our simulation model are outlined in **Appendix Table 1.** Key assumptions implemented in the model are listed in **Appendix Table 2.**

#### *Population projection*

The population projections stratified by sex and age for Belgium were obtained from Statbel, the Belgian Statistical Office.[28] As Statbel does not have the population projections by education level (low, middle, high), we assumed that the relative difference in population estimates across education levels by age and sex for the simulation period from 2022 to 2041 was equal to the relative differences from the census in 2021.[29] We defined educational level as follows: low (from no education to lower secondary education), middle (upper secondary and post-secondary non-tertiary education), and high (from short-cycle tertiary education to doctoral degree level education). In Belgium, we excluded individuals with "unknown" and "not applicable" information on educational status. Therefore, our estimated impacts of the policy may be underestimated.

The German population projections by sex and age were derived from the German Federal Statistical Office.[30] The population projections do not have stratification by educational level and we used population size and composition data from 2013-2019 to estimate relative differences by educational level.[31] We applied these differences to the population projections for the simulation period from 2022 to 2041. We defined educational level as described above for Belgium.

#### *Cardiovascular disease (CVD) mortality projection*

Using the "demography" package,[32] we projected mortality trends to 2041, by age, sex, and education levels, based on the number of annual CVD deaths observed from 2012 to 2020 by Statbel for Belgium (*data provided upon request to Statbel*). For Germany, we projected mortality trends by age and sex using data from the German Information System of the Federal Health Monitoring (*Gesundheitsberichterstattung des Bundes*) based on annual CVD deaths from 1991 to 2019.[20,33] CVD death counts include coronary heart disease (CHD) (ICD-10: I20 to I25) and overall strokes (ICD-10: I60 to I69, I64, I69.4, and I69.8). Our mortality projection based on previous data would account for potential continuing declines in CVD mortality. This approach helps to avoid overestimating the benefits of any CVD intervention.[1] As the mortality projection for Germany was not stratified by education level, our simulation model incorporated information on sex- and education-specific relative risk (RR) from a previous study[34] to simulate CVD mortality by education level.

#### *Body mass index (BMI)*

Our estimates for the exposures (BMI, energy and SSB intakes) used data from nationally representative surveys: National Food Consumption Survey (FCS) 2014-2015 for Belgium,[35,36] and Cooperative Health Research in the Region Augsburg (*Kooperative Gesundheitsforschung in der Region Augsburg*) (KORA) S4, F4, FF4 (1999, 2007, 2014) and German National Nutrition Survey (*Nationale Verzehrstudie*) (NVS) II (2006) for

Germany.[37,38] As we only used single-year survey data for Belgium (2014), we did not model any trends of the exposures.

For both country, we used generalised additive models for location, shape and scale (GAMLSS) ("gamlss" package[39]), flexible models that can handle complex relationships between different types of variables,[40,41] to estimate the distribution of BMI. GAMLSS created all parameters of an assumed distribution of BMI conditional on some function of some variables or predictors, such as year (for Germany only), age, sex and education level (for both Belgium and Germany). All the parameters of the assumed BMI distribution were applied to the population projections throughout the simulation period (2022 – 2041) to estimate BMI.

#### *Out-of-home energy intake*

Due to the absence of out-of-home energy intake data in both countries, and as we do not have information on the frequency of out-of-home consumption in either Belgium or Germany, we calculated out-of-home energy intake by multiplying overall daily energy intake with the proportions of daily out-of-home energy intake reported by previous studies based on national survey data in 2004 in Belgium[42] and data collected in 2000 in two study centres (cities) in Germany.[43]

We used GAMLSS to create the parameters of the distribution of daily energy intake conditional on year (for Germany only), age, sex and education level (for both Belgium and Germany). We then applied the parameters to the population projections to estimate overall daily energy intake. To estimate out-of-home energy intake, we then multiplying overall daily energy intake from GAMLSS with the proportions of daily out-of-home energy intake specific by age group and sex.

These estimations of the out-of-home energy intake have some limitations. The study used in Belgium to assess the proportions of daily out-of-home energy intake also considered eating in a friend's house as eating out-of-home,[42] and the study used for Germany may not be nationally representative as the data collected in two study centres only.[43] In addition, we did not estimate out-of-home energy intake by educational level as this information was not available in either study (only by age and sex). Finally, we assumed that the proportions of energy intake from eating out have remained stable since early 2000's. We may underestimate the effect of the policy (mandatory menu calorie labelling) as eating out might be more common due to changes in food environments.[44,45]

#### *Non-diet SSB intake*

We estimated overall SSB intake throughout the simulation period based on GAMLSS parameters of an assumed distribution of SSB intake conditional on year (for Germany only), age, sex and education level (for both Belgium and Germany). To calculate non-diet SSB intake, we multiplied overall SSB intake with the proportions of non-diet SSB intake by age and sex in Germany created using GAMLSS based on KORA FF4 (2014) study.[20] In Belgium, as almost all the participants from FCS 2014-2015 were consumers of non-alcoholic sugary drinks (99-100%), we assumed that all SSB intake was from non-diet SSBs.

### Estimating the effect of change in energy and SSB intake on BMI and CVD mortality

#### *Estimating the effect of change in energy intake on BMI*

Following a previous approach,[1] we calculated the reduction in energy intake (in kcal) due to the implementation of mandatory menu calorie labelling by subtracting the level of energy intake post-intervention from baseline intake for each year. We assumed that menu calorie labelling would immediately affect energy intake, and this effect would remain consistent throughout the simulation horizon (2022 – 2041).

Changes in energy intake would have a subsequent immediate impact on BMI. To transform a change in energy intake into an equivalent change in body weight, we used a formula developed by Christiansen & Garby[46] based on energy conservation principles:

$$\Delta BW = k * \Delta\left(\frac{Energy\ intake}{Physical\ activity\ level}\right)$$

Change in body weight ($\Delta BW$) is in kilogram (kg) and energy intake is in MegaJoule (MJ). Physical activity level (PAL) is computed as the total energy expenditure divided by the resting energy expenditure. A constant value ($k$)

is calculated based on both fundamental principles of energy conservation and directly measured data (constant values of 17.7 and 20.7 are assigned for men and women, respectively).[46]

We assumed that the policy has no impact on physical activity levels, and therefore, PAL was kept constant at 1.5 [46] to represent limited physical activity.[47] We calculated the equivalent change in BMI based on the estimated change in body weight assuming constant individuals' height.

***Estimating the effect of change in BMI on CVD mortality***

The increased risk of CVD mortality for one standard deviation (SD) increase in BMI (4.56 kg/m²) for those with a BMI ≥ 20 kg/m² was informed by the Emerging Risk Factors Collaboration (ERFC).[48] A risk of 1 was assigned for individuals with a BMI < 20 kg/m², while the risks for other individuals with a BMI ≥ 20 kg/m² were determined based on sex- and smoking-adjusted age-specific estimates for CVD mortality from the ERFC [48], taking into account the new change in BMI.

We calculated the population-attributable risk fraction (PARF) which represents the proportion of CVD mortality attributable to a specific risk factor (BMI ≥ 20 kg/m²). In microsimulation modelling where individuals have different risks due to their risk factor (BMI) and characteristics (e.g., age), PARF can be calculated as follows (see [49] for detailed information including mortality calculation).

$$PARF = \frac{n}{\sum_{i=1}^{n} RR_{BMI,i}} \quad [\mathbf{1}]$$

$n$ refers to the total number of (synthetic) individuals in the simulation modelling and $RR_{BMI}$ represents the unique individual relative risk of CVD mortality due to BMI as the risk factor.

We then calculated the proportion of CVD mortality not attributable to the risk factor using the following formula.

$$M_{Theoretical\ minimum} = M_{Observed} \text{ x } (1 - PARF) \quad [\mathbf{2}]$$

$M_{Theoretical\ minimum}$ is the estimated CVD mortality if the risk factor is optimal, derived from multiplying the observed CVD mortality ($M_{Observed}$) and the proportion not attributable to the risk factor (1- PARF). Assuming the PARF is consistent after the initial (or baseline) year, $M_{Theoretical\ minimum}$ is calculated by age, sex, and SES for all years of the simulation period.

Finally, we can calculate the individualised annual probability of CVD mortality due to their risk factor (BMI) and other varying characteristics (e.g., age, sex, SES) by assuming that $M_{Theoretical\ minimum}$ is the annual baseline probability of CVD mortality not because the modelled risk factors (other risk factors than BMI).

$$P(CVD|age, sex, SES, BMI) = M_{Theoretical\ minimum} \text{ x } \left(RR_{BMI,i}\right) \quad [\mathbf{3}]$$

We used the formulas above for CHD and stroke separately and then calculated CVD mortality as the sum of CHD and stroke. We calculated the number of CVD deaths prevented or postponed (DPPs) by subtracting the total number of CVD deaths in a policy scenario from the total number of CVD deaths in the counterfactual scenario. For each different scenario, we present corresponding aggregated CVD DPPs across the simulation period. It is important to note that, we assumed no lag time between energy intake and BMI as the change in energy intake has an immediate effect (< one year) on BMI. However, we used a 5-year lag time (e.g., as in [50]) for the impact of the change in BMI on CVD mortality risk. Therefore, the policy has no impact on CVD mortality in the first 5 years of implementation (2022-2026), but it impacts the population from 2027 up to the simulation period to 2041.

***Estimating the effect of change in SSB intake on BMI and CVD mortality***

We assumed that a change in SSB intake will have simultaneous impact on CVD mortality through BMI (indirect effect) and without BMI (direct effect). Therefore, our main findings consider both indirect and direct effects of SSB intake on CVD mortality.

To calculate the indirect effect of change in SSB intake on CVD mortality, through BMI, we used BMI-specific estimates from a meta-analysis of three prospective cohorts by Micha et al.[51] Assuming linearity, a decrease in one serving of SSB (~ 227.3045 mL) will lead to a decrease in BMI of 0.10kg/m$^2$ (0.05–0.15) in individuals with BMI < 25 kg/m$^2$ and of 0.23 kg/m$^2$ (0.14–0.32) in individuals with BMI ≥ 25 kg/m$^2$. We assumed the immediate effect of change in SSB intake on change in BMI (no lag time). The change in BMI due to SSB intake was then transformed into the equivalent change or increase in CVD mortality risk using the estimates from the ERFC [48] as described above.

To calculate the direct effect, we also used an estimate from Micha et al.[51] that calculated age-specific BMI-adjusted relative risk of one SSB serving per day on CVD mortality from four cohort studies. Similar to mandatory menu calorie labelling, we assumed aa lag time of 5 years between exposure (SSB intake) and the outcome (CVD mortality risk) (e.g., as in [50]).

Using the estimated individual post-policy CVD mortality risk due to the decreased SSB intake through both pathways, we estimated PARF, CVD mortality, and CVD DPPs for each SSB tax scenario using the approach described above for the mandatory menu calorie labelling policy (see [49]). We also present CVD DPPs-related SSB intake estimated through BMI pathway alone (indirect effect) as part of the sensitivity analyses.

### Estimating model uncertainty

The Monte Carlo approach[52] with 200 iterations was used to estimate the uncertainty from different model parameters incorporated in the simulation modelling. There are different potential sources of uncertainty, including the sampling errors of baseline data, the uncertainty of GAMLSS parameters to predict BMI, energy, and SSB intakes by age, sex, and SES (see "Creating synthetic population"), mortality forecasts, the relative risk of BMI on the outcomes (CHD, stroke), and the uncertainty of the assumed policy (menu calorie labelling, SSB) effects.

### Appendix Table 1. Data sources used in the model

| Parameters | Outcome | Details | Differences by sociodemographic groups | Source | Projection distribution of the mean | Uncertainty |
|---|---|---|---|---|---|---|
| ***Population data*** | | | | | | |
| Population projection | Population | Belgium: Population projection 1992-2071<br>Germany: Population projection 2019-2060 | Stratified by year, age, sex | Belgium: Statbel[28]<br>Germany: The Federal Statistical Office [30] | - | Population projection |
| Population size and composition | Population | Belgium: Census 2021<br>Germany: Official population data 2013-2019 | Stratified by age, sex, education level | Belgium: Statbel[29]<br>Germany: The Federal Statistical Office [31] | - | - |
| Mortality | Deaths from CVD (CHD, stroke) | Belgium: CVD deaths 2012- 2020<br>Germany: CVD deaths 1991-2019 | Stratified by year, age, sex, education (in Belgium only), cause of death | Belgium: Statbel (*data provided upon request)*<br>Germany: German Information System of the Federal Health Monitoring[33] Information on sex- and education-specific RR from a previous study[34] was incorporated to simulate CVD mortality by education level. | Log normal | Mean ± SD |
| ***Exposures*** | | | | | | |
| BMI | BMI | Belgium: FCS 2014-2015<br>Germany: KORA S4, F4, FF4 (1999, 2007, 2014) and NVS II (2006) for Germany | Stratified by year (in Germany only), age, sex, education | Belgium: FCS 2014-2015[35,36]<br>Germany: KORA S4, F4, FF4 (1999, 2007, 2014) and NVS II (2006) for Germany[37,38] | GAMLSS | GAMLSS parameters |
| Energy intake | Energy intake | Belgium: FCS 2014-2015<br>Germany: KORA S4, F4, FF4 (1999, 2007, 2014) and NVS II (2006) for Germany | Stratified by year (in Germany only), age, sex, education | Belgium: FCS 2014-2015[35,36]<br>Germany: KORA S4, F4, FF4 (1999, 2007, 2014) and NVS II (2006) for Germany[37,38] | GAMLSS | GAMLSS parameters |
| SSB intake | SSB intake | Belgium: FCS 2014-2015<br>Germany: KORA S4, F4, FF4 (1999, 2007, 2014) and NVS II (2006) for Germany | Stratified by year (in Germany only), age, sex, education | Belgium: FCS 2014-2015[35,36]<br>Germany: KORA S4, F4, FF4 (1999, 2007, 2014) and NVS II (2006) for Germany[37,38] | GAMLSS | GAMLSS parameters |
| Proportions of out-of-home energy intake | Energy out-of-home | Belgium: Based on national survey data in 2004<br>Germany: Based on data collected in two study centres in 2000 | Stratified by age and sex | Belgium: Vandevijvere et al.[42]<br>Germany: Orfanos et al.[43] | - | - |

| | | | | | | |
|---|---|---|---|---|---|---|
| Proportions of non-diet SSB | Non-diet SSB | Belgium: Assuming all SSB intake from non-diet SSB based on FCS 2014-2015<br>Germany: KORA FF4 (2014) | (in Germany only) Stratified by age and sex | Belgium: FCS 2014-2015[35,36]<br>Germany: KORA FF4 (2014)[38] | GAMLSS | GAMLSS parameters |
| ***Effect estimates*** | | | | | | |
| Effect of menu calorie labelling | Change in energy intake | An estimate from a meta-analysis of 19 intervention studies and randomised control trials | No differential effect | Shangguan et al.[9] | - | Mean ± SD |
| Effect of menu calorie labelling on product reformulation | Change in energy intake | An estimate reported in the US following the implementation of menu calorie labelling policy | No differential effect | Bleich et al.[15-17], Du et al.[8] | - | Mean |
| Effect of SSB tax | Change in SSB intake | An estimate of demand price elasticity (i.e., % change in sales or consumption due to % change in price) and a pass-through rate (i.e., the extent of increase in price passed on to customers) from a meta-analysis of 33-41 studies | No differential effect | Andreyeva et al.[22] | - | Mean ± SD |
| Effect of SSB tax on product reformulation | Change in sugar intake | Findings from studies on the impact of the soft drinks industry levy (SDIL) in the UK on a reduction in the sugar content of all SSB products sold. | No differential effect | von Philipsborn et al.,[25] Bandy et al.,[26] Emmert-Fees et al.[20] | - | |
| Effect of change in energy intake on BMI | Change in BMI | A formula based on energy conservation principles | Stratified by sex | Christiansen & Garby[46] | - | - |
| Effect of change in SSB or sugar intake on BMI | Change in BMI | An estimate from a meta-analysis of three prospective cohorts | Stratified by age and baseline BMI | Micha et al.[51] | Log normal | Mean ± SD |
| Effect of change in BMI on CVD mortality risk | Change in CVD mortality | A collaborative analysis of 58 prospective studies | Stratified by age and baseline BMI | Emerging Risk Factors Collaboration[48] | Log normal | Mean ± SD |
| Effect of change in SSB or sugar intake CVD mortality risk | Change in CVD mortality | A BMI-adjusted estimate from a meta-analysis of four prospective cohorts | Stratified by age | Micha et al.[51] | Log normal | Mean ± SD |

BMI = body mass index; CHD = coronary heart disease; CVD = cardiovascular disease; FCS = Food Consumption Survey; GAMLSS = generalised additive models for location, shape and scale; KORA = Cooperative Health Research in the Region Augsburg (*Kooperative Gesundheitsforschung in der Region Augsburg*); NVS = German National Nutrition Survey (*Nationale Verzehrstudie*); Statbel = Belgian Statistical Office; SSB = sugar-sweetened beverage; SD = standard deviation; RR = relative risk

### Appendix Table 2. Assumptions implemented in the model

| Components | Key assumptions |
|---|---|
| Population data | We do not consider social mobility (i.e., individuals remain at the same educational level) and therefore, the population composition by educational level is stable throughout the simulation period. |
| Exposures | The surveys on which exposures were based were truly representative of the population. |
| | The distribution of exposures by sex, age, and education for which we did not include time trends (e.g., in Belgium) remains the same over the simulation period. |
| | Energy and SSB purchased are equivalent to intake (or consumption) |
| | The proportion of out-of-home large businesses corresponds to the proportion of out-of-home calories consumed from these businesses (for mandatory menu calorie labelling). |
| Effect estimates | There are no differential effects of the policies across sociodemographic characteristics (age, sex, education level). |
| | The effect of the policies on intake or consumption remains stable over time. |
| | We assume multiplicative risk effects. |

### Appendix Table 3. Estimates of baseline population, out-of-home energy, SSB intake, and obesity status (in 2022)

| Characteristics | Belgium | Germany |
|---|---|---|
| Population size estimate (aged 30-89) (*total)* | 7,010,188 | 58,050,700 |
| Low education | 2,463,001 (35.13%) | 6,928,617 (11.94%) |
| Middle education | 2,306,847 (32.91%) | 32,393,488 (55.80%) |
| High education | 2,240,340 (32.96%) | 18,728,595 (32.26%) |
| Out-of-home energy intake estimate (kcal) *(mean; median)* | 408.60; 347.78 | 475.61 ; 420.62 |
| Low education | 348.64 ; 268.04 | 458.54 ; 395.27 |
| Middle education | 440.97 ; 377.77 | 454.23 ; 396.92 |
| High education | 441.20 ; 386.40 | 518.81 ; 468.20 |
| Non-diet SSB intake estimate (ml) *(mean; median)* | 109.03 ; 0 | 79.04 ; 15.15 |
| Low education | 116.94 ; 0 | 117.00 ; 18.50 |
| Middle education | 138.12 ; 0 | 73.60 ; 14.60 |
| High education | 70.36 ; 0 | 74.41 ; 15.10 |
| Obesity status estimate (≥ 30 kg/m$^2$) *(%)* | 27.38% | 18.44% |
| Low education | 33.77% | 24.90% |
| Middle education | 30.96% | 19.75% |
| High education | 17.78% | 13.79% |

**Appendix Table 4. Ratios of DPP rates (per 100,000 population) between low- and high-education groups for mandatory menu calorie labelling and the SSB tax, based on combined consumer response and reformulation scenarios**

| Scenarios | Belgium | | Germany | |
|---|---|---|---|---|
| | **Ratio** | **Probability of Ratio > 1** | **Ratio** | **Probability of Ratio > 1** |
| **Mandatory menu calorie labelling** | | | | |
| Partial implementation | 0.00 | 0.00 | 0.00 | 0.33 |
| Full implementation | 0.86 | 0.49 | 0.76 | 0.29 |
| **SSB taxes** | | | | |
| 10% | 2.59 | 0.90 | 1.91 | 0.87 |
| 20% | 2.59 | 0.91 | 1.91 | 0.89 |
| 30% | 3.31 | 0.91 | 2.00 | 0.91 |

A ratio (in median) of > 1 indicates greater rates of DPPs in low than high education groups.

### Appendix Table 5. Sensitivity analyses for mandatory menu calorie labelling using minimum (11%) and maximum compensation (42%)

| Scenarios | Belgium | | Germany | |
|---|---|---|---|---|
| | **Changes in obesity prevalence** | **CVD DPPs** | **Changes in obesity prevalence** | **CVD DPPs** |
| | | **Minimum compensation (11%)** | | |
| ***Consumer response*** | | | | |
| Partial implementation | -0.07 (-0.11, -0.04) | 30[a] (0, 200) | -0.27 (-0.39, -0.17) | 1500 (0, 5000) |
| Full implementation | -2.33 (-3.26, -1.29) | 1000 (200, 2600) | -2.79 (-3.81, -1.95) | 18000 (5500, 38000) |
| ***Combined*** | | | | |
| Partial implementation | -0.13 (-0.18, -0.09) | 59[a] (0, 400) | -0.49 (-0.61, -0.38) | 2800 (500, 7000) |
| Full implementation | -3.96 (-4.76, -2.99) | 1600 (400, 3800) | -4.67 (-5.56, -3.94) | 32000 (12000, 64000) |
| | | **Maximum compensation (42%)** | | |
| ***Consumer response*** | | | | |
| Partial implementation | -0.05 (-0.08, -0.02) | 24[a] (0, 200) | -0.18 (-0.26, -0.11) | 1000 (0, 4000) |
| Full implementation | -1.57 (-2.11, -0.88) | 600 (0, 1800) | -1.89 (-2.59, -1.28) | 12000 (3000, 26000) |
| ***Combined*** | | | | |
| Partial implementation | -0.11 (-0.14, -0.08) | 43[a] (0, 400) | -0.39 (-0.48, -0.32) | 2500 (500, 6000) |
| Full implementation | -3.23 (-3.85, -2.58) | 1400 (200, 3600) | -3.88 (-4.54, -3.35) | 26000 (9000, 51000) |

Estimates are presented for 20 years from the policy implementation (2022 to 2041) with the population-level impacts observed from 2027 to 2041 due to a 5-year lag time.

[a]Estimates are presented as mean because the median is 0 (zero).

Estimates are presented as median and 95% uncertainty intervals (UIs), unless otherwise specified.

CVD = cardiovascular disease; DPPs = Deaths prevented or postponed

**Appendix Table 6. Sensitivity analyses for mandatory menu calorie labelling using percentages of turnover of large out-of-home businesses**

| Scenarios | Belgium | | Germany | |
|---|---|---|---|---|
| | **Changes in obesity prevalence** | **CVD DPPs** | **Changes in obesity prevalence** | **CVD DPPs** |
| | **Percentages of turnover (Belgium = 10% ; Germany = 21%)** | | | |
| ***Consumer response*** | | | | |
| Partial implementation | -0.20 (-0.30, -0.11) | 90[a] (0, 400) | -0.52 (-0.74, -0.34) | 3000 (500, 8000) |
| ***Combined*** | | | | |
| Partial implementation | -0.39 (-0.49, -0.29) | 200 (0, 610) | -1.00 (-1.23, -0.82) | 6000 (1500, 13000) |

Estimates are presented for 20 years from the policy implementation (2022 to 2041) with the population-level impacts observed from 2027 to 2041 due to a 5-year lag time.

[a]Estimates are presented as mean because the median is 0 (zero).

Estimates are presented as median and 95% uncertainty intervals (UIs), unless otherwise specified.

CVD = cardiovascular disease; DPPs = Deaths prevented or postponed

### Appendix Table 7. Sensitivity analyses for SSB tax using different effects reported in other meta-analyses

| Scenarios | **Belgium** | | **Germany** | |
|---|---|---|---|---|
| | **Changes in obesity prevalence** | **CVD DPPs** | **Changes in obesity prevalence** | **CVD DPPs** |
| | **Effects from different meta-analyses** | | | |
| ***Consumer response*** | | | | |
| 10% tax (Afsin et al.) | -0.03 (-0.07, -0.01) | 400 (0, 1000) | -0.03 (-0.06, -0.01) | 2500 (0, 6000) |
| 10% tax (Teng et al.) | -0.05 (-0.09, -0.02) | 400 (0, 1600) | -0.05 (-0.09, -0.01) | 3500 (500, 9000) |
| ***Combined*** | | | | |
| 10% tax (Afsin et al.) | -0.17 (-0.26, -0.11) | 1400 (400, 4000) | -0.17 (-0.23, -0.11) | 12000 (4000, 20000) |
| 10% tax (Teng et al.) | -0.18 (-0.28, -0.12) | 1600 (400, 4200) | -0.17 (-0.25, -0.11) | 12000 (4500, 22000) |

Estimates are presented for 20 years from the policy implementation (2022 to 2041) with the population-level impacts observed from 2027 to 2041 due to a 5-year lag time.
Estimates are presented as median and 95% uncertainty intervals (UIs), unless otherwise specified.
CVD = cardiovascular disease; DPPs = Deaths prevented or postponed

### Appendix Table 8. Sensitivity analyses for the indirect effect of SSB tax on CVD mortality through BMI only

| Scenarios | Belgium | Germany |
|---|---|---|
| | **CVD DPPs** | **CVD DPPs** |
| ***Consumer response*** | | |
| 10% tax | 33[a] (0, 200) | 280[a] (0, 1000) |
| 20% tax | 64[a] (0, 400) | 500 (0, 2000) |
| 30% tax | 100[a] (0, 600) | 500 (0, 2500) |
| ***Reformulation*** | | |
| 30% decrease in sugar | 83[a] (0, 410) | 500 (0, 2000) |
| ***Combined*** | | |
| 10% tax | 99[a] (0, 600) | 500 (0, 2500) |
| 20% tax | 130[a] (0, 600) | 1000 (0, 3500) |
| 30% tax | 150[a] (0, 610) | 1000 (0, 3500) |

Estimates are presented for 20 years from the policy implementation (2022 to 2041) with the population-level impacts observed from 2027 to 2041 due to a 5-year lag time.
[a]Estimates are presented as mean because the median is 0 (zero).
Estimates are presented as median and 95% uncertainty intervals (UIs), unless otherwise specified.
CVD = cardiovascular disease; DPPs = Deaths prevented or postponed
